# A deep intronic variant in *MME* causes autosomal recessive Charcot-Marie-Tooth neuropathy through aberrant splicing

**DOI:** 10.1101/2024.04.22.24306048

**Authors:** Bianca R Grosz, Jevin M Parmar, Melina Ellis, Samantha Bryen, Cas Simons, Andre L.M. Reis, Igor Stevanovski, Ira W. Deveson, Garth Nicholson, Nigel Laing, Mathew Wallis, Gianina Ravenscroft, Kishore R. Kumar, Steve Vucic, Marina L Kennerson

## Abstract

**Background:** Loss-of-function variants in *MME* (membrane metalloendopeptidase) are a known cause of recessive Charcot-Marie-Tooth Neuropathy (CMT). A deep intronic variant, *MME* c.1188+428A>G (NM_000902.5), was identified through whole genome sequencing (WGS) of two Australian families with recessive inheritance of axonal CMT using the seqr platform. *MME* c.1188+428A>G was detected in a homozygous state in Family 1, and in a compound heterozygous state with a known pathogenic *MME* variant (c.467del; p.Pro156Leufs*14) in Family 2.

**Aims:** We aimed to determine the pathogenicity of the *MME* c.1188+428A>G variant through segregation and splicing analysis.

**Methods:** The splicing impact of the deep intronic *MME* variant c.1188+428A>G was assessed using an *in vitro* exon-trapping assay.

**Results:** The exon-trapping assay demonstrated that the *MME* c.1188+428A>G variant created a novel splice donor site resulting in the inclusion of an 83 bp pseudoexon between *MME* exons 12 and 13. The incorporation of the pseudoexon into *MME* transcript is predicted to lead to a coding frameshift and premature termination codon (PTC) in *MME* exon 14 (p.Ala397ProfsTer47). This PTC is likely to result in nonsense mediated decay (NMD) of *MME* transcript leading to a pathogenic loss-of-function.

**Interpretation:** To our knowledge, this is the first report of a pathogenic deep intronic *MME* variant causing CMT. This is of significance as deep intronic variants are missed using whole exome sequencing screening methods. Individuals with CMT should be reassessed for deep intronic variants, with splicing impacts being considered in relation to the potential pathogenicity of variants.

## Introduction

Charcot-Marie-Tooth neuropathy (CMT) is the most common inherited peripheral neuropathy, affecting 1/2500 individuals^1^. CMT is characterized by progressive length-dependent loss of peripheral motor and sensory nerves, resulting in distal muscle weakness and sensory symptoms^2^. Patients are broadly divided into subtypes based on whether nerve conduction studies (NCS) indicating demyelinating (CMT1) or axonal (CMT2) forms of the disease. Recessive loss-of-function variants in the membrane metalloendopeptidase (*MME*) gene have been previously reported to cause axonal Charcot-Marie-Tooth neuropathy type 2T (CMT2T; OMIM: #617017)^3–11^. *MME* encodes for neprilysin, a widely expressed membrane-bound metallopeptidase that has a key role in neuropeptide processing^12^. A significant portion of patients with axonal CMT remain genetically undiagnosed^13–17^, indicating that further disease-causing genes and pathogenic variants in known genes are yet to be identified.

Splicing variants are increasingly being recognized as a cause of Mendelian disease^18,19^. The precise removal of introns (non-coding regions) and inclusion of exons (coding regions) in the final mature mRNA relies on the spliceosome and auxiliary splicing factors recognizing specific sequence motifs, such as the 5’ donor splice site and 3’ acceptor splice site. Splice-altering variants can weaken or abolish recognition of the correct splice sites, or alternatively strengthen or create cryptic splice sites that mimic consensus splicing sequences^20^. These variants typically lead to one or more mis-splicing events that result in the skipping of partial or complete exons, and/or the retention of partial or complete introns ^21–42^. Pathogenic splicing variants have been found in several CMT genes including *MPZ*^21,30,39,40^, *MFN2*^22,30,31^, *LRSAM1*^23^, *IGHMBP2*^24^, *INF2*^25^, *MCM3AP*^26^, *SH3TC2*^30,32,38^*, GDAP1*^27,42^, *SBF1*^28^, *NDRG1*^37^, and *FGD4*^29^. Variants affecting canonical splice donor and acceptor sites have also been described in *MME* ^4,41^.

Exon-trapping, also known as a mini-gene assay, is an *in vitro* technique used to identify exons in a genomic region of interest^43^. This is of particular use when relevant patient tissue is unavailable or when relevant transcripts may be unstable or degraded by nonsense mediated decay (NMD) ^44^. The genomic region of interest is cloned into an exon-trapping vector between two known exons. This exon-trapping vector is transfected into a cell line where it is transcribed and undergoes a series of post-transcriptional processes that include pre-mRNA splicing to create mature mRNA. This mRNA consists of the ‘trapped’ exons of the genomic region of interest flanked by the known exons, which can then be Sanger sequenced to characterize the ‘trapped’ exons. Comparison of ‘trapped’ exons between wild type and variant genomic sequences can also indicate if a candidate variant affects splicing ^45–51^. The well-validated exon-trapping vector pSpliceExpress ^52^ consists of known exons of the rat insulin gene, *Ins2*, and has been used previously to determine the splicing impacts of multiple pathogenic variants in Mendelian disease ^45–51^.

Here we report a deep intronic variant in *MME* [chr3:155142758A>G (hg38); *MME* c.1188+428A>G], found in a recessive state in two Australian families. Exon-trapping revealed that this variant creates a novel splice donor site in *MME* intron 12 (NM_000902.5) which, along with a preceding existing cryptic splice acceptor site, results in the incorporation of an 83 bp pseudoexon in the *MME* transcript [chr3:155,142,675-155,142,757 (hg38); r.1188_1189ins[1188+345_1188+427]]. The coding frameshift caused by this pseudoexon leads to a PTC in exon 14 and likely NMD of the *MME* transcript (p.Ala397ProfsTer47), resulting in a loss of *MME* function.

## Materials and Methods

### Subjects

Members of Family 1 were recruited and informed consent was obtained for this study using protocols approved by the Sydney Local Health District Human Ethics Research Committee (2019/ETH07839). Recruitment and informed consent for the proband of Family 2 was approved by the Human Research Ethics Committee of the Royal Melbourne Hospital (HREC/16/MH/251).

### Variant Detection

Genomic DNA for Family 1 was extracted from peripheral blood using the PureGene Kit (Qiagen) following the manufacturer’s instructions. WGS for two individuals in Family 1 (V:1 and V:3) was outsourced to the Garvan Sequencing Platform. Paired-end sequencing reads of 150 base pairs were generated using the Illumina NovaSeq 6000 sequencing machine, with 30-fold average read depth.

In Family 2, genomic DNA for the proband was extracted at the Department of Diagnostic Genomics (PathWest, Perth, Australia) using the QIAsymphony^SP^ machine and QIAsymphone® DSP DNA Midi Kit. The proband underwent WGS at the Australian Genomics Research Facility (AGRF), Melbourne, following GATK4 best-practices. Paired-end sequencing reads of 150 base pairs were generated using the Illumina NovaSeq 6000 sequencing machine, with 30-fold average read depth. Parental DNA was not available for sequencing.

WGS data processing was performed at the Centre for Population Genomics (CPG) following the DRAGEN GATK best practices pipeline. Reads were aligned to the hg38 reference genome using Dragmap (v1.3.0). Cohort-wide joint calling of single nucleotide variants (SNVs) and small insertion/deletion (indel) variants was performed using GATK HaplotypeCaller (v4.2.6.1) with “--dragen-mode” enabled. Sample sex and relatedness quality checks were performed using Somalier (v0.2.15)^53^. Variants were annotated using VEP 105, and loaded into the web-based variant filtration platform, seqr^54^. A WGS search was conducted using the seqr^45^ platform for low minor allele frequency (<0.01) variants in CMT-related genes, for both Family 1 and 2 [gene list-Hereditary Neuropathy_CMT_IsolatedAndComplex (Version 2.14)^55^]. Additionally, variants were analyzed by the CPG Automated Interpretation Pipeline (AIP, https://github.com/populationgenomics/automated-interpretation-pipeline). *In silico* splicing analysis was conducted using SpliceAI ^56^.

### Segregation Analysis

The *MME* c.1188+428A>G variant in Family 1 and Family 2 was amplified using primers that spanned *MME* intron 12 (5’-CTCAGCCGAACCTACAAGGA-3’; 5’-GCAAATGCTGCTTCCACAT-3’) to produce a 1264 bp amplicon [chr3:155,142,289-155,143,552 (hg38)]. An internal sequencing primer was used (5’-CTGTGTTAAAAGTAATTTCGGGG-3’) and the amplicon was Sanger sequenced. The *MME* c.467del variant in Family 2 was amplified (5’-GCAGAGCCGTATGCATCACT-3’; 5’-TTCAGCTGTCCAAGAAGCACC-3’). A 717bp amplicon was produced [chr3:155,116,171-155,116,887 (hg38)], which was subsequently Sanger sequenced.

### Sanger Sequencing

For Family 1, PCR amplicons were sent to Garvan Molecular Genetics, Garvan Institute (Sydney, Australia) for Sanger sequencing using BigDye Terminator cycle sequencing protocols. For Family 2, proband PCR amplicons were Sanger sequenced at AGRF, Perth using BigDye Terminator sequencing protocols. Sequences were visualized and analyzed using Snapgene Software version 7.1 (www.snapgene.com).

### Oxford Nanopore Technologies (ONT) Long Read Sequencing

High molecular weight (HMW) DNA samples of the Proband in Family 2 were transferred to the Garvan Sequencing Platform for targeted long-read sequencing analysis on ONT instruments. Prior to ONT library preparations, DNA was sheared to ∼20-25 kb fragment size using a MegaRuptor 3 instrument and visualized post-shearing on an Agilent FemtoPulse.

Sequencing libraries were prepared from ∼3-5 µg of HMW DNA, using native library prep kit SQK-LSK114, according to manufacturer’s instructions. Each library was loaded onto a R10.4.1 flow cell and sequenced on a PromethION device with live target selection/rejection executed by the ReadFish software package^57^. Detailed descriptions of software and hardware configurations used for ReadFish are provided in a previous publication^58^. Samples were run for a maximum duration of 72 h, with nuclease flushes and library reloading performed at approximately 24- and 48-h timepoints for targeted sequencing runs, to maximize sequencing yield. Raw ONT sequencing data was converted to BLOW5 format^59^ using slow5tools (v.0.3.0)^60^ then base-called using Guppy (v6). Resulting FASTQ files were aligned to the hg38 reference genome using minimap2 (v2.14-r883)^61^. Variants were called using clair3^62^, phased using Whatshap^63^ and visualized using the Integrative Genomics Viewer (IGV, v2.17.3)^64^.

### Cell Culture

The human HeLa cervical epithelial cell line (ATCC) was maintained in Dulbecco’s Modified Eagle’s Medium (DMEM)(Gibco) containing 10% (v/v) fetal bovine serum (FBS) (Gibco), +100 U/mL penicillin (Gibco), and 100 μg/mL streptomycin (Gibco) at 37°C in humidified air and 5% CO2.

### Cloning Procedures

The region surrounding *MME* c.1188+428A>G [chr3:155,141,790-155,143,755 (hg38)] was amplified from the genomic DNA of a heterozygous carrier of the c.1188+428A>G variant, using *attB* adapter primers (5’-AAAAAGCAGGCTTCGCTCTTAAATGGTTGGCTT-3’; 5’- AGAAAGCTGGGTAACTAGACTCTTGGGGAAGGC -3’). The *MME* amplicon was then cloned into the exon-trapping pSpliceExpress vector between flanking *Ins2* exons using a two-step Gateway cloning BP reaction (ThermoFisher). pSpliceExpress was a gift from Stefan Stamm (Addgene plasmid #32485)^52^. The pSpliceExpress-*MME* clones were then Sanger sequenced to verify the correct insertion of *MME* and to determine the c.1188+428A>G genotype of each clone (as the genomic DNA template was heterozygous for the variant).

### *In vitro* exon-trapping

HeLa cells were grown to approximately 70% confluence in a 6-well plate. HeLa cells were separately transfected with either 2 μg of pSpliceExpress-*MME*_WT_ or 2 μg of pSpliceExpress- *MME*_c.1188+428A>G_ using Lipofectamine 3000 (ThermoFisher). RNA extraction was performed 48 h following transfection using the RNEasy Mini Kit (Qiagen), and reverse-transcribed template was prepared using the iScript cDNA Synthesis Kit (Bio-Rad). PCR amplification of cDNA was conducted using primers designed to anneal to the flanking *Ins2* exons (5’- CAGCACCTTTGTGGTTCTCA-3’; 5’-CAGTGCCAAGGTCTGAAGGT-3’). The RT-PCR amplicons were size fractionated using a 1.5% w/v agarose gel. The largest amplicon for each vector was gel-purified using Isolate II PCR and Gel Kit (Bioline) for Sanger Sequencing.

### *In silico* splicing analysis of reported *MME* variants

All reported *MME* variants in gnomAD v.4.0.0 were detected by searching the gnomAD browser for the genomic region corresponding with the *MME* gene; ‘3-155024124-155183704’(hg38). The *MME* variants reported in gnomAD were then exported using the ‘Export Variants to CSV’ function. The consequences of the variants were described by gnomAD using the Variant Effect Predictor (VEP) annotation based on the most deleterious predicted functional effect of each variant ^65^ . These variants were then filtered for a minor allele frequency (MAF) <0.01 and a maximum SpliceAI Δscore >0.8 (high precision splicing change prediction^66^) as annotated by gnomAD.

## Results

### Clinical Phenotypes

Family 1 consists of four affected siblings from a consanguineous family of European (non-Finnish) background (Figure 1a). The phenotype was consistent with a generalized sensorimotor axonal neuropathy and sensory ataxia without cerebellar signs (Table 1), and was confirmed with NCS for individuals V:1 and V:6 (Table 2). Affected individuals had previously undergone diagnostic and research whole exome sequencing with negative results.

**Figure 1:**
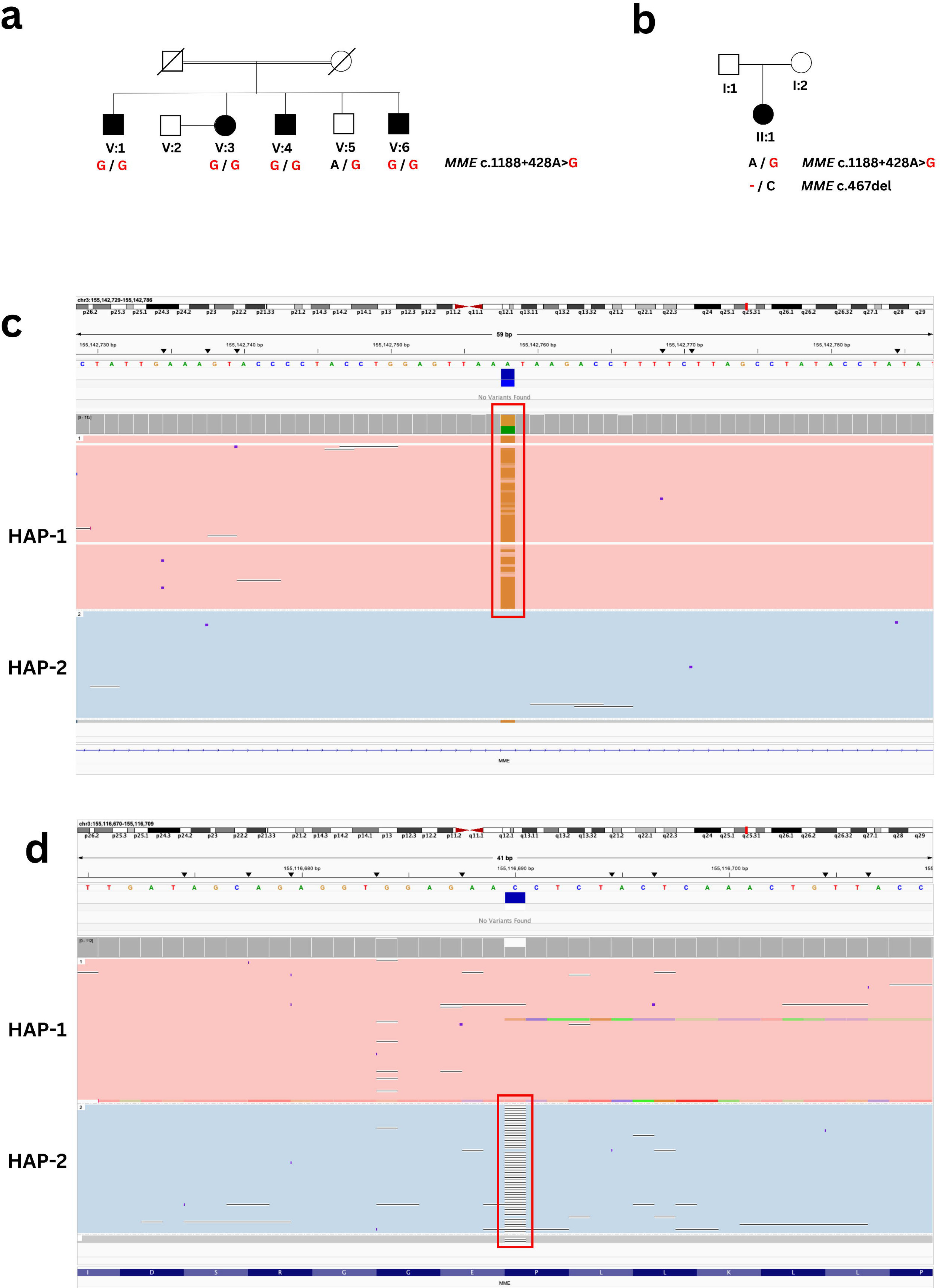
The *MME* c.1188+428A>G (NM_000902.5) variant segregates with recessive CMT in two families. A) Family 1 pedigree showing the associated genotypes for the *MME* c.1188+428A>G (NM_000902.5) variant. Affected individuals in the fifth generation are homozygous (G/G). Unaffected individuals in the fifth (V) and sixth (VI) generation are heterozygous (A/G). Squares represent males and circles represent females, solid symbol denotes affected individual. The double line in the fourth (IV) generation indicates a consanguineous relationship. The *MME* c.1188+428 genotype is denoted beneath individuals who underwent Sanger sequencing, with the pathogenic ‘G’ allele in red text. B) A pedigree showing the associated genotypes for *MME* c.1188+428A>G and *MME* c.467del in the index individual in Family 2. C-D) Haplotype phasing showed that the c.1188+428A>G (C) and c.467del (D) variants were present on alternative haplotypes (light red and light blue), thus inherited *in trans* in the proband of Family 2. Phased targeted ONT long-read sequencing reads were visualized using the Integrative Genomics Viewer (IGV; v2.17.3).

**Table 1.** Phenotypic characterization of individuals with recessive CMT *MME* variants reported in this manuscript. Abbreviations-AFO: Ankle foot orthoses; CMTNS: Charcot-Marie-Tooth Neuropathy Score; UL/LL: Upper Limb/Lower Limb.

| Family # | Family 1 |  |  |  | Family 2 |
| --- | --- | --- | --- | --- | --- |
| Individual | V:1 | V:3 | V:4 | V:6 | II:1 |
| Sex | M | F | M | M | F |
| Onset | Fourth decade | Childhood | Childhood | Childhood | Early-adult |
| Age at evaluation<br>(years) | 60-65 | 65-70 | 65-70 | 65-70 | 50-55 |
| Presenting symptom | Gait unsteadiness | Difficulty running | Difficulty sitting on crossed legs during childhood | Difficulty running, multiple ankle sprains | Bilateral leg weakness and sensory disturbance |
| Atrophy UL/LL | Yes/yes | Yes/yes | Yes/yes | Yes/yes | No/Yes<br>(left calf only) |
| Foot deformity | None | Pes cavus/hammer toes | - | Pes cavus | Pes cavus |
| Tone UL/LL | Normal/reduced | Normal/normal | Normal, increased |  | Normal/normal |
| Shoulder abduction | 5 | 5 | 3 | 5 | 4 |
| Elbow flexion | 4 | 5 | 3 | 4 | 5 |
| Elbow extension | 4 | 5 | 3 | 4 | 5 |
| Finger abduction | 2 | 4 | 3 | 1 | 4 |
| Hip flexion | 4 | 3 | 3 | 3 | 4 |
| Knee extension | 4 | 3 | 3 | 4 | 4 |
| Knee flexion | 4 | 3 | 3 | 4 | 4 |
| Ankle dorsiflexion | 0 | 0 | 3 | 1 | 3 |
| Ankle plantarflexion | 0 | 0 | 3 | 0 | 4 |
| Deep tendon reflexes UL/LL | Absent/absent | Absent ankle | Hyperreflexia knee, absent ankle | Absent/absent | Normal/ reduced ankle reflexes |
| Plantars | Absent | - | Absent | Flexor | Flexor |
| Proprioception UL/LL | Normal/reduced to the MTP | - | - | Normal/reduced to the ankle | Normal/reduced to ankle |
| Vibration UL/LL | Normal/reduced to ankle | Absent in toes | - | Reduced to wrist/reduced to knees | Reduced to mid-shin |
| Pinprick UL/LL | Normal/reduced to ankle | Normal/reduced to the mid shin | Reduced to elbows/reduced to knees | Reduced to PIP joint/reduced to knees | Reduced to mid-shin<br>Reduced dorsum hand |
| Temperature sensation UL/LL | Normal/reduced to ankle | - | - | Reduced to wrist/reduced to above knee | Reduced to mid-shin |
| Other symptoms/signs | Cough | GORD and cough | Corticospinal tract signs | Nil | Fatty infiltration on left gastrocnemius |
| Gait | High steppage/ataxic | High steppage/ataxic | Steppage gait | High steppage/ataxic | High steppage |
| Romberg's sign | Positive | Positive |  | Positive | Mild sway |
| Mobility aids | AFO, walking stick and mobility scooter | AFO, walker | AFO, walker, mobility scooter | Wheelchair | AFO, elbow crutch |
| CMTNS (version 2) | 31 | - | 28/36 | 36 | 15 |

**Table 2.** Nerve conduction study (NCS) findings of individuals with recessive CMT *MME* variants reported in this manuscript. Abbreviations: APB: abductor pollicis brevis; ADM: abductor digiti minimi; EDB: extensor hallucis brevis; FHB: flexor hallus brevis; CMAP: compound motor action potential; CV: conduction velocity; NR: no response; SNAP: sensory nerve action potential; R: right, L:left.

| Family # | Family 1 |  | Family 2 |
| --- | --- | --- | --- |
| Individual | V:1 | V:6 | II:1 |
| Age at NCS (years) | 60-65 | 65-70 | 55-60 |
| Median nerve (digit II) [orthodromic(R)] |  |  |  |
| SNAP (uV) | NR | NR | 6.6 |
| CV (m/s) | NR | NR | 50 |
| Ulnar nerve (digit V) [orthodromic(R)] |  |  |  |
| SNAP (uV) | NR | 1 | 4.9 |
| CV (m/s) | NR | 40.2 | 51 |
| Sural nerve [orthodromic(R)] |  |  |  |
| SNAP (uV) | NR | NR | NR |
| CV (m/s) | NR | NR | NR |
| Sural nerve [orthodromic(L)] |  |  |  |
| SNAP (uV) | NR | NR | NR |
| CV (m/s) | NR | NR | NR |
| Median nerve (APB) (R) |  |  |  |
| CMAP (mV) | NR | NR | 4.8 |
| CV (m/s) | NR | NR | 41 |
| Ulnar nerve (ADM) (R) |  |  |  |
| CMAP (mV) | 2 | 0.6 | 6.6 |
| CV (m/s) | 44.4 | 44.7 | 52 (wrist-below elbow)<br>57 (wrist-above elbow) |
| Peroneal nerve (EDB) (R) |  |  |  |
| CMAP (mV) | NR | NR | NR |
| CV (m/s) | NR | NR | NR |
| Tibial nerve (FHB) (L) |  |  |  |
| CMAP (mV) | NR | NR | NR |
| CV (m/s) | NR | NR | NR |
| Needle EMG | Absent voluntary units in the right tibialis anterior and medial gastrocnemius muscles, as well as denervation-reinnervation changes in | Absent recruitment of motor units in the right tibialis anterior, medial gastrocnemius, first dorsal interosseous and vastus lateralis | Reduced recruitment of motor units in the right tibialis anterior and right vastus medialis muscles. Discrete recruitment of muscle fibres in the |
|  | the right vastus lateralis muscle. | muscles. Denervation-reinnervation changes were evident in the right deltoid, biceps brachii and triceps brachii muscles. | right gastrocnemius muscle. |

Family 2 consists of one affected female born to a healthy, non-consanguineous couple (Figure 1b). Neurological examination revealed mild distal upper limb weakness and moderate distal lower limb weakness (Table 1). NCS showed evidence of an axonal sensorimotor neuropathy (Table 2). Bilateral MRI of the thighs and calves showed muscle atrophy (Supplementary Figure 1).

### Genetic Analysis

WGS screening using the seqr platform in Family 1 revealed a single homozygous variant in two affected individuals (V:1 and V:3), *MME* c.1188+428A>G (NM_000902.5). This variant was reported in dbSNP build 155^67^ (rs61758195), with a low MAF in gnomAD v4.0.0^68^ (10/152120), All of Us (35/490,748)^69^, and TOPMED^70^ (15/264290). No homozygous individuals were reported in gnomAD, All of Us, or TOPMED. SpliceAI predicted that *MME* c.1188+428A>G could create a strong novel splice donor site (Score 0.97; where a SpliceAI score above 0.8 is considered a ‘high precision’ predicted splice variant^56^). This in turn strengthened the SpliceAI prediction for a cryptic splice acceptor site 83 bp upstream of the novel splice donor site (Score 0.99).

Segregation analysis in Family 1 confirmed the biallelic inheritance of the *MME* c.1188+428A>G variant segregated with the CMT phenotype (Figure 1a). Sanger sequencing of DNA from available individuals showed that all affected individuals were homozygous for the *MME* c.1188+428A>G variant and unaffected individuals were carriers (Supplementary Figure 2).

WGS screening using the seqr platform and AIP was conducted in the proband of Family 2 (II:1). The *MME* c.1188+428A>G variant was detected in a compound heterozygous state with a second *MME* variant (*MME* c.467del; p.Pro156Leufs*14), which has previously been reported as pathogenic^41^ (Figure 1b). Sanger sequencing validated the presence of each *MME* variant in the index individual (Supplementary Figure 3). As parental DNA was unavailable, ONT long-read sequencing was conducted to phase the heterozygous *MME* variants in the proband (Figure 1c-d). ONT long-read sequencing confirmed that *MME* c.1188+428A>G (boxed red in Figure 1c) and c.467del (boxed red in Figure 1d) were present on alternative haplotypes (hap-1: red, hap-2: blue) and therefore were in *trans* in the proband.

### *In vitro* exon-trapping of *MME*-pSpliceExpress vectors

Two separate exon-trapping pSpliceExpress vectors were generated to assess the *in vitro* splicing impact of the *MME* c.1188+428A>G variant: pSpliceExpress-*MME*_WT_ (wild-type) and pSpliceExpress-*MME*_c.1188+428A>G_ (variant). A schematic of the constructs and relevant SpliceAI scores are shown in Figure 2a. RT-PCR products produced following transfection of these vectors were analyzed using gel electrophoresis (Figure 2b), which revealed a visible size difference between the wild-type (381 bp) and *MME* c.1188+428A>G amplicons (464 bp). The gel-purified amplicons were Sanger sequenced and the sequencing was aligned to the WT *MME* mRNA sequence. The sequenced products showed that the pSpliceExpress-*MME*_WT_ produced a transcript that was correctly spliced between *MME* exon 12 and 13 (Figure 2c). In contrast, exon-trapping of the pSpliceExpress-*MME*_c.1188+428A>G_ vector showed that an 83 bp pseudoexon had been spliced between *MME* exon 12 and 13 (Figure 2d). A BLAT search^71^ using the sequence of the trapped pseudoexon revealed alignment to the intronic region directly upstream of the *MME* c.1188+428A>G variant [chr3:155,142,675-155,142,757 (hg38)]. This suggests that the *MME* c.1188+428A>G variant creates a novel splice donor site leading to the aberrant inclusion of 83 bp of intronic *MME* sequence in the final spliced transcript, as predicted by SpliceAI. Prediction of the novel *MME* coding sequence caused by the introduction of the pseudoexon showed that a PTC was generated in exon 14 (p.Ala397ProfsTer47) at genomic position chr3:155,144,368 (hg38)(Supplementary Figure 4). This PTC likely leads to NMD of the *MME* transcript.

**Figure 2.**
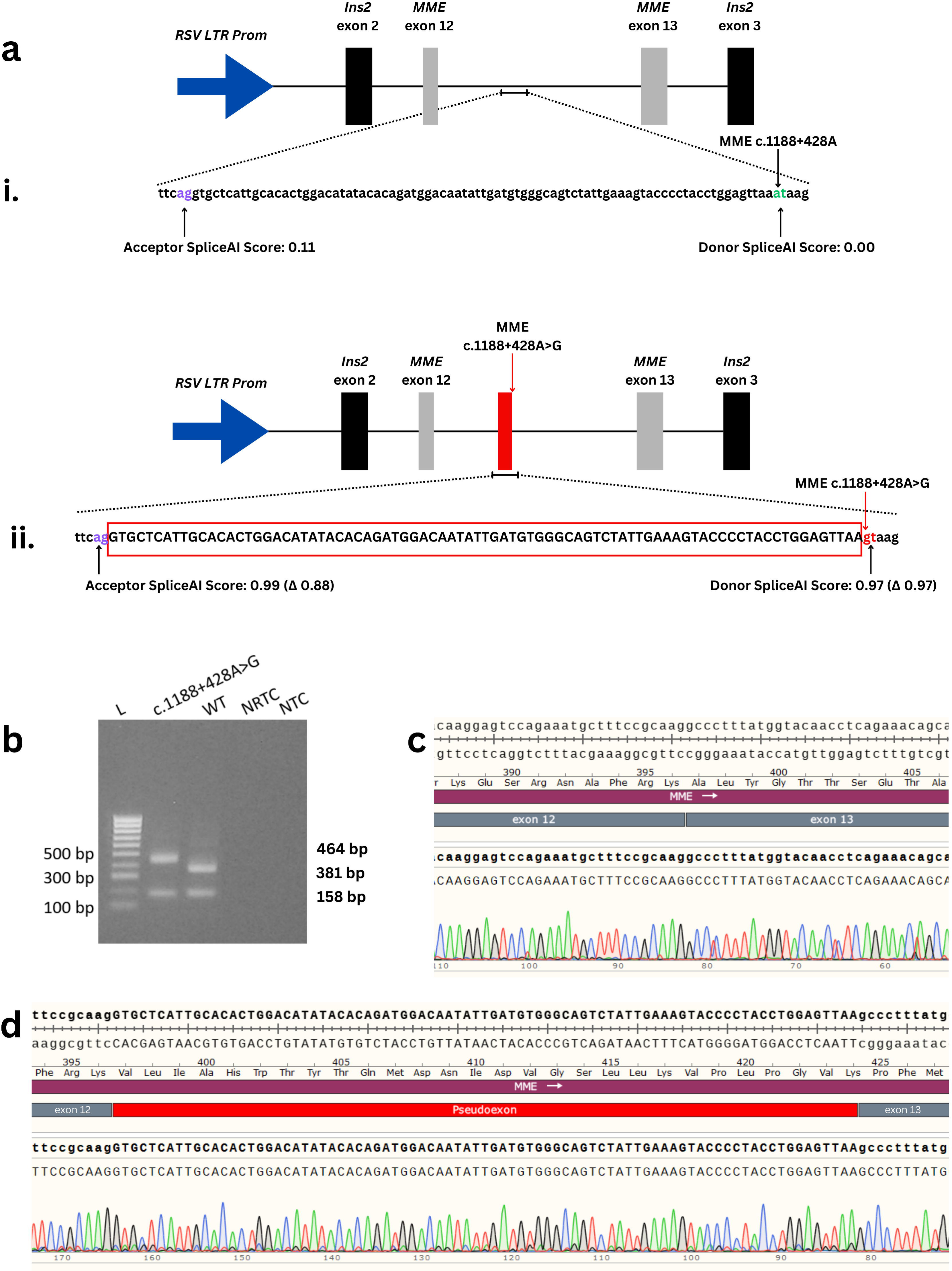
An exon-trapping assay was used to analyze splicing changes caused by the *MME* c.1188+428A>G variant. A) Schematic of the pSpliceExpress-*MME*_WT_ (wild-type) and pSpliceExpress-*MME*_c.1188+428A>G_ (variant) constructs. Both constructs consisted of an RSV LTR promoter region (blue) controlling transcription of a minigene of *MME* exons 12 and 13 (grey) flanked by *Ins2* exon 2 and *Ins2* exon 3 (black). The constructs differed in the presence of either an A (wild type: green text) or a G allele (mutant: red text) at *MME* c.1188+428A>G. i) The wild type sequence was not predicted to contain any strong splice sites when assessed using SpliceAI, with a score of 0.11 for the acceptor site (purple text) and 0.00 for the donor site (green text). ii) SpliceAI predicted that the *MME* c.1188+428A>G variant (red arrow) would create a strong splice donor site [Score: 0.97 (Δ 0.97); red text], which then strengthened the prediction of a splice acceptor site 83 bp upstream [Score 0.99 (Δ0.97); purple text]. The predicted 83 bp pseudoexon sequence is capitalized and boxed in red. B) The amplicon produced by the pSpliceExpress-*MME*_c.1188+428A>G_ vector (464 bp) showed a visible increase in product size when compared to the amplicon produced by pSpliceExpress-*MME*_WT_ (381 bp). The 158 bp RT-PCR product produced by both vectors indicated splicing and ligation of the flanking *Ins2* exon 2 and exon 3. Lanes: L: HyperLadder 100 bp (Bioline); WT: wild-type; NRTC: negative cDNA conversion control (no reverse transcriptase). NTC: negative PCR reaction control (no cDNA template). C) Sanger sequencing of the pSpliceExpress-*MME*_WT_ RT-PCR product confirmed correct splicing between *MME* exon 12 and 13. D) Sanger sequencing of the pSpliceExpress-*MME*_c.1188+428A>G_ RT-PCR product revealed the presence of a novel 83 bp pseudoexon (red) between *MME* exon 12 and 13. Abbreviations: RSV LTR: Rous Sarcoma Virus Long Terminal Repeat promoter; *Ins2*: rat preproinsulin 2 gene.

### *In silico* splicing analysis of *MME* variants

All reported *MME* variants in gnomAD v.4.0.0 were assessed using the integrated SpliceAI scores to determine if splicing variants in *MME* were a likely underrecognized cause of disease. There were 37,264 total variants reported in *MME* in gnomAD, of which 35,673 variants had a MAF <0.01. Of these, 88 variants had a maximum SpliceAI Δscore above 0.8 (Supplementary Table 1). There were no homozygotes reported for any of the 88 *MME* putative splicing variants. The majority of these predicted splicing variants were predicted to directly change either the canonical splice donor sites (26/88) or canonical splice acceptor sites (30/88) of *MME*, including an inframe deletion (c.1317_1317+2del) and a frameshift variant (c.957+1del). An additional nine variants were predicted to affect a splicing region, including one which is also annotated as a synonymous variant (c.1188G>A; p.Lys396Lys). Nine missense variants (p.Asp209Gly, p.Ile217Ser, p.Glu282Val, p.Arg365Ile, p.Ser436Gly, p.Asp533Gly, p.Ile553Val, p.Val554Phe, p.Gln692Arg) and a synonymous variant (p.Gly417Gly) were also predicted to alter splicing. Twelve variants were annotated as ‘intron variants’, of which two could be considered ‘deep intronic’ variants (c.1188+428A>G, described in this manuscript, and c.197-9871A>G). Interestingly, further analysis using SpliceAI demonstrated that *MME* c.197-9871A>G was predicted to create a splice donor site (Score 0.81) and strengthen an upstream cryptic splice acceptor site (Score 0.81) in a similar manner to c.1188+428A>G, thereby possibly creating an in-frame 96 bp pseudoexon.

## Discussion

Here we report a deep intronic variant, *MME* c.1188+428A>G causing recessive CMT2T in two unrelated Australian families. This variant results in a pseudoexon that likely leads to NMD of the *MME* transcript, resulting in a loss-of-function. This is in keeping with previously reported *MME* variants which have broadly been characterised as ‘loss-of-function’ variants. To our knowledge, this is the first report of a pathogenic deep intronic variant in *MME*. Given that a significant portion of patients with axonal CMT remain genetically undiagnosed^13–17^, it’s possible that deep intronic variants in *MME* explain a portion of this diagnostic gap.

Individuals from Family 1 previously underwent both diagnostic WES and research WES, and the proband in Family 2 previously underwent targeted gene panel testing (PathWest neuro v3). These testing methods did not capture deep intronic regions and returned negative results. However, deep intronic regions have previously been shown to have a higher prevalence of variants than coding regions and canonical splice sites^72^. Our findings suggest that intronic SNPs should be analyzed to determine if they impact splicing before they are dismissed as benign. Detection and functional validation of deep intronic variants has previously been shown to increase diagnostic rates in other Mendelian diseases including X-linked Alport syndrome^73^, inherited retinal disorders^74,75^, and dystrophinopathy^76,77^. Pathogenic deep intronic variants, such as described here, are likely to be underreported amongst CMT-causing genes due to a lack of detection by WES and targeted gene panels, and a lack of functional investigation upon detection.

The *MME* c.1188+428A>G variant was reported in multiple different genetic ancestry groups in gnomAD v.4.0.0^68^, including in the European (Non-Finnish) (8/68008), African/African American (1/41432), and ‘remaining’ (1/2092) ancestry groups. This was also reflected in the ‘All of Us’ Research Program^69^, which reported the *MME* c.1188+428A>G variant in the African (2/107,888) and European populations (33/256,804). Whilst it is possible that *MME* c.1188+428A>G represents a recurrent *de novo* variant, it is also likely that this variant has persisted at low levels in the global population. As this variant is missed by WES and may not be prioritised by variant-filtering approaches focusing on coding variants, this variant may therefore represent an underappreciated cause of recessive CMT. This is further supported by its detection in two Australian CMT families who are not known to be related.

Whilst the predicted splicing variants reported in *MME* are individually rare, we have described eighty-eight variants in gnomAD that are predicted by SpliceAI to alter splicing. Nine of these variants were annotated as missense variants, and one was a synonymous change. This is of note as these ‘missense’ variants are often assumed to result in a single amino acid change, and synonymous variants are often considered functionally neutral, with their effect on splicing typically not assessed^78^. Our results suggest that the discovery of any of these eighty-eight variants in a homozygous or compound heterozygous state in an individual with CMT should prompt further functional investigation of their effect on splicing of the *MME* transcript.

Prior to functional validation, *MME* c.1188+428A>G was considered a variant of uncertain significance (VUS) according to American College of Medical Genetics and Genomics (ACMG) criteria^79^ (BP4, PM2, PM3, PP1), However, the functional evidence generated by the splicing assay now allows for the addition of PVS1 (null variant in a gene where loss-of-function is a known mechanism of disease) and PS3 (well-established *in vitro* or *in vivo* functional studies supportive of a damaging effect on the gene or gene product) criteria. This allows reclassification of the variant as ‘pathogenic’. Therefore, this work demonstrates the importance of functional validation to confirm the effect of candidate variants on splicing to increase diagnostic rates for those with inherited disease.

Previously described homozygous *MME* patients typically have a phenotype consistent with late-onset axonal neuropathy. In contrast, three of the four affected siblings in Family 1 described childhood onset and the proband of Family 2 described symptom onset in early adulthood. It has also previously been reported that heterozygous *MME* variants can cause spinocerebellar ataxia type 43 (SCA43)^80^ as well as autosomal dominant CMT^3^. However, the individuals in Family 1 who were *MME* c.1188+428A>G heterozygotes were all clinically assessed as neurologically normal, including an individual who is in their seventh decade. Homozygous affected individuals in Family 1 were noted to have a sensory ataxia rather than cerebellar ataxia, although MRI brain studies were not conducted. Therefore, our findings here do not support a role for *MME* c.1188+428A>G to cause SCA43 or autosomal dominant CMT, and further expand the phenotype of recessive CMT2T.

Understanding the specific effects of splice-affecting variants is crucial for developing potential therapeutic strategies. Antisense oligonucleotides (ASOs) that modulate splicing are an active area of research, including individualized approaches to treat rare genetic diseases^81–83^. Several antisense nucleotides that modify splicing have been approved by the United States Food and Drug Administration and have resulted in marked improvements in clinical outcomes in those with genetic diseases^84–94^. An FDA-approved “n-of-1” ASO, milasen, successfully blocked pathogenic pseudoexon inclusion in *MFSD8* in a patient with neuronal ceroid lipofuscinosis type 7^94^. ASOs which block pathogenic pseudoexons, such as that created by *MME* c.1188+428A>G, have also been described in *in vivo* preclinical models^93,95–99^. As such, individuals with the *MME* c.1188+428A>G variant may represent a form of CMT that is treatable through personalized ASO therapy and warrants further investigation.

## Data Availability

All data produced in the present study are available upon reasonable request to the authors.

## Acknowledgements

This work was funded by the Medical Research Future Fund (MRFF) Genomics Health Futures Mission (APP2007681). JMP is supported by the Australian Government Research Training Program. IWD was supported by MRF2025138 & MRF2023126. Genomic analysis was supported by the Centre for Population Genomics (Garvan Institute of Medical Research and Murdoch Children’s Research Institute) and was funded in part by a National Health and Medical Research Council investigator grant (2009982) and the Medical Research Future Fund (MRFF) Genomics Health Futures Mission (2008820).

