## Supplementary for "A deep intronic variant in *MME* causes autosomal recessive Charcot-Marie-Tooth neuropathy through aberrant splicing"

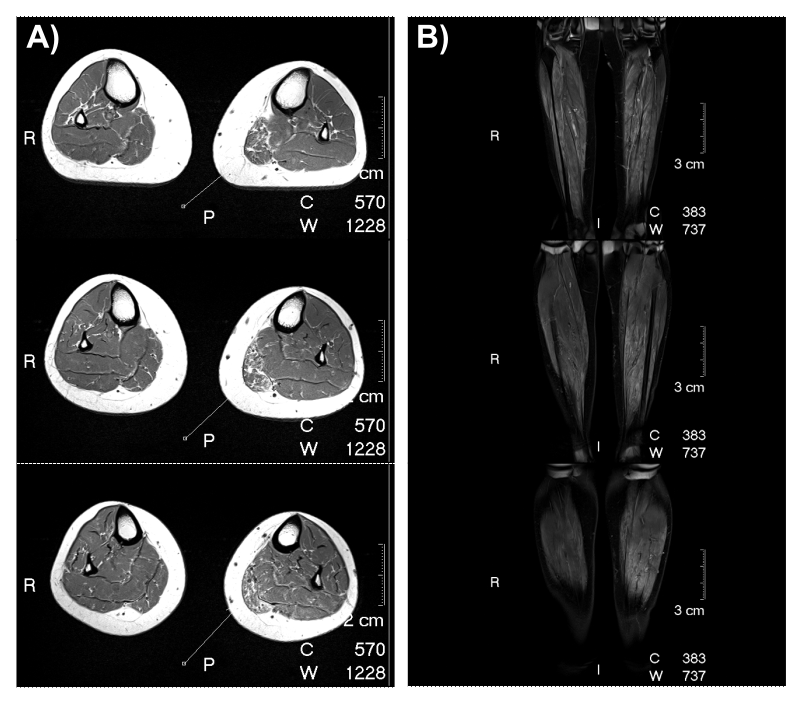
Supplementary Figure 1: MRI images of the proband from Family 2**.** Bilateral transverse (A) and coronal (B) MRI of the calf muscles showing multifocal calf muscle T2 hyperintensity consistent with myositis (left > right). MRI shows focal left medial gastrocnemius abnormality including atrophy and T1 hyperintense fatty infiltration. Transverse MRI images (A) are arranged proximal to distal and Coronal MRI images (B) are arranged anterior to posterior.

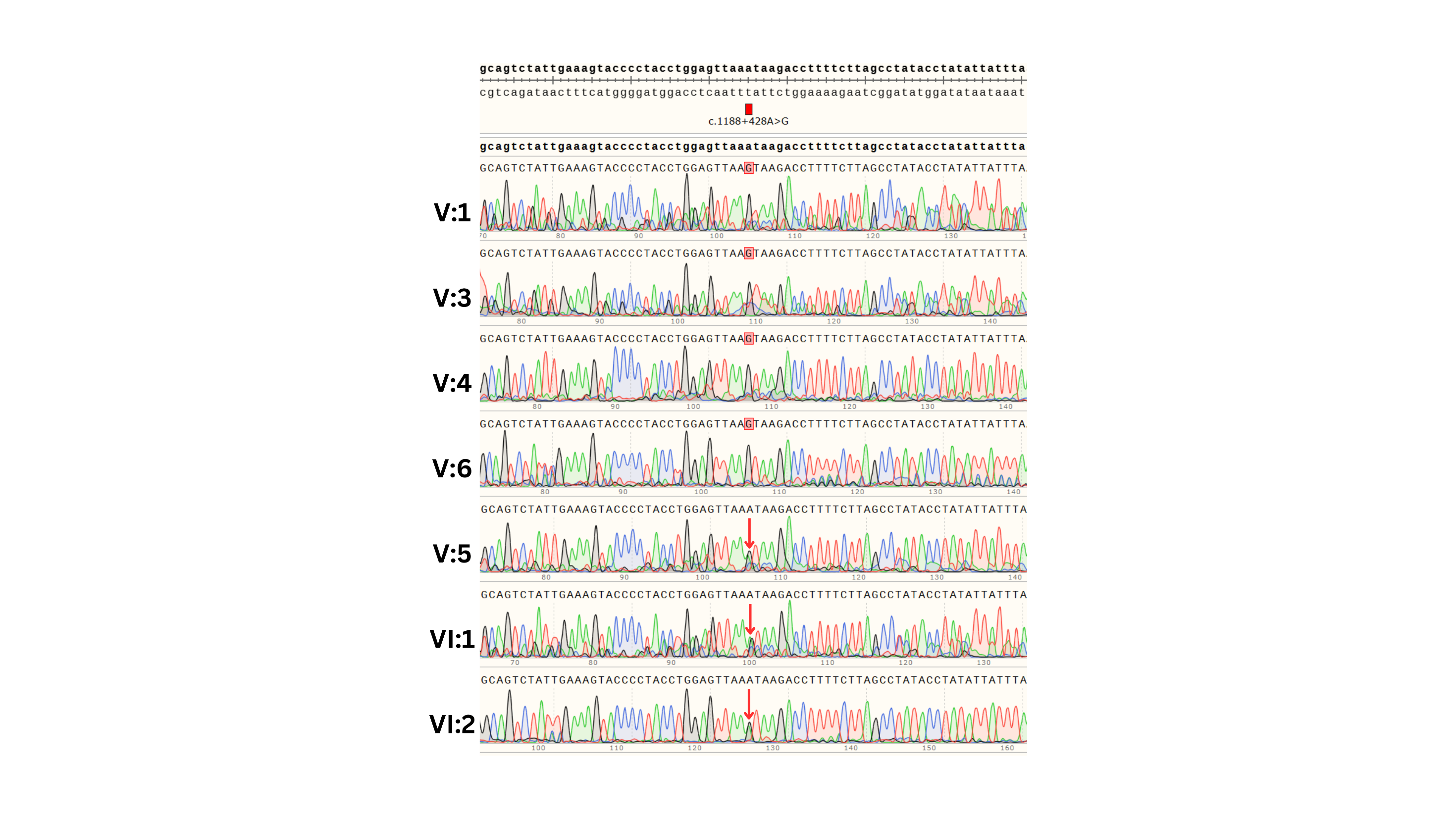
Supplementary Figure 2: Sequence traces of affected homozygous individuals (V:1, V:3, V:4, V:6) and unaffected heterozygous individuals (V:5, VI:1, V1:2) from Family 1. The position of the *MME* c.1188+428A>G variant is indicated by a red box for the homozygous individuals and an arrow for the heterozygous individuals.

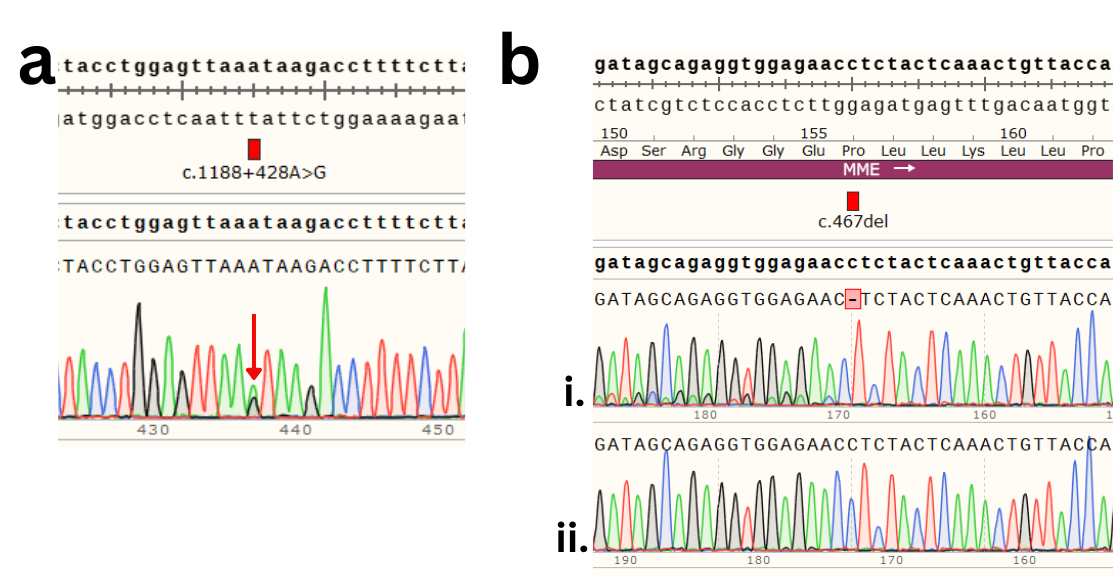

Supplementary Figure 3: Sanger sequencing of the proband of Family 2 (II:1) confirmed heterozygosity for both *MME* variants, c.1188+428A>G and c.467del. a) Sequence trace showing heterozygosity for the c.1188+428A>G variant (red arrow). b) Sequence trace of the proband (i) showing heterozygosity for the c.467del variant (red box). The sequencing primer was in the reverse orientation and the base deletion results in heterozygous reads for the bases upstream^1^. The difference in peak sizes is likely due to previous reported allelic dropout in this region A control individual (ii) is shown for comparison.

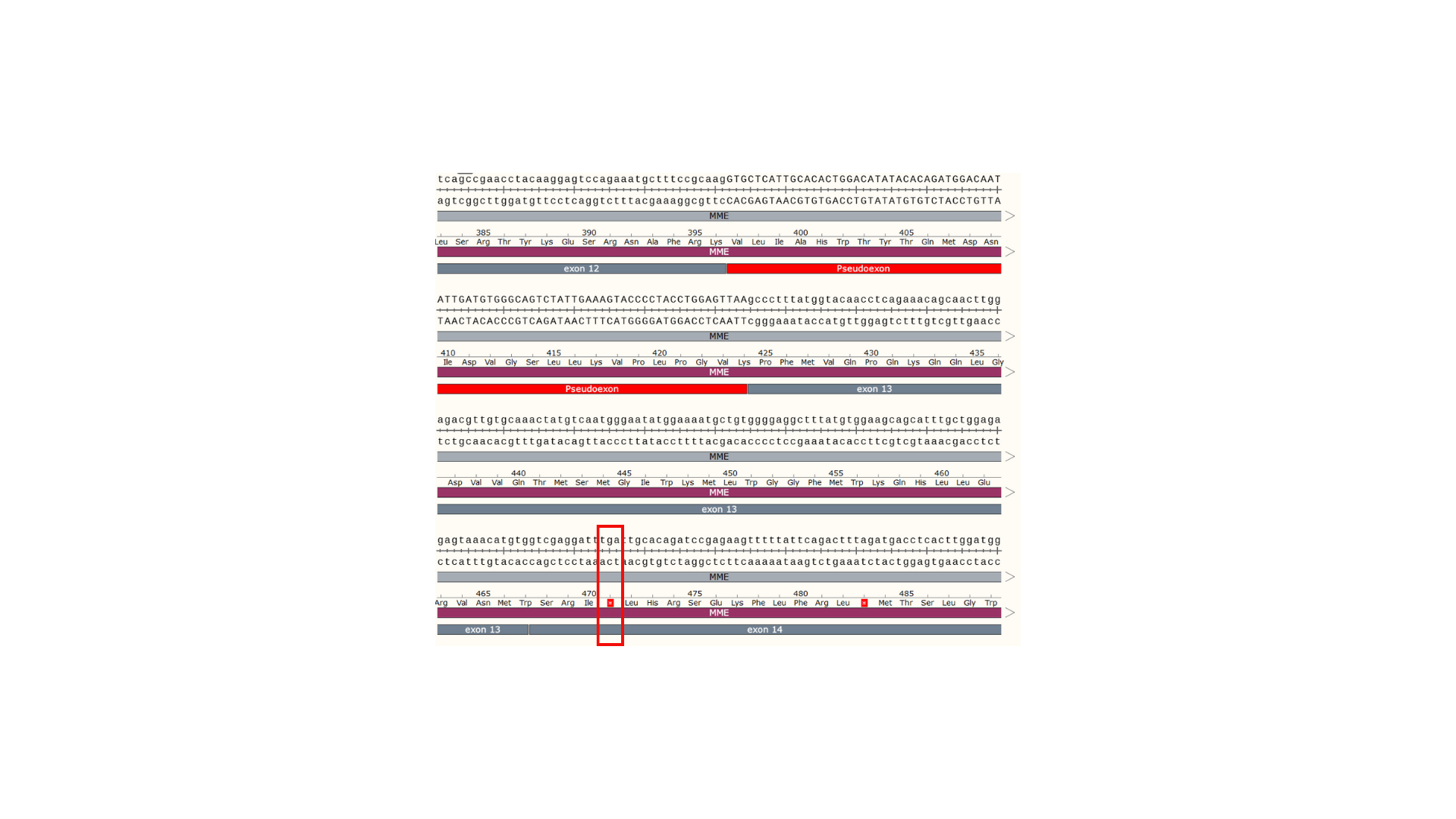

Supplementary Figure 4: Analysis of the novel coding sequence produced by the incorporation of the 83 bp pseudoexon (red) into the *MME* transcript shows a coding frameshift is introduced resulting in a premature termination codon (PTC) in *MME* exon 14 (p.Ala397ProfsTer47; PTC boxed red and indicated by a red asterisk).

Supplementary Table 1: Variants in *MME* reported in gnomAD (v4.0.0)^2^ that are predicted to cause a splicing change when assessed by SpliceAI. Abbreviations: VUS: Variant of Uncertain Significance; LP: Likely Pathogenic; P: Pathogenic; IV: intron variant; SAV: splice acceptor variant; SD: splice donor variant; MV: missense variant; HGVS: Human Genome Variation Society; SRV; splice region variant; FS: frameshift variant; SynV: synonymous variant; InFD: inframe deletion; VEP: Variant Effect Prediction; MAF: minor allele frequency.

| **gnomAD ID [chr-base(hg38)-ref-alt]** | **rsIDs** | **Transcript** | **HGVS Consequence** | **VEP** | **ClinVar** | **MAF** | **SpliceAI max ∆score** |
| --- | --- | --- | --- | --- | --- | --- | --- |
| 3-155024329-G-A | rs1368004215 | ENST00000492661.5 | c.-11+5G>A | IV |  | 3/152210 | 0.84 |
| 3-155084156-A-G | rs1034739692 | ENST00000360490.7 | c.-10-2A>G | SAV |  | 1/628378 | 1 |
| 3-155084157-G-C | rs1085307704 | ENST00000360490.7 | c.-10-1G>C | SAV |  | 2/1460418 | 1 |
| 3-155084157-G-T | rs1085307704 | ENST00000360490.7 | c.-10-1G>T | SAV | VUS | 1/1460416 | 1 |
| 3-155084327-GGTGA-G | rs1345316322 | ENST00000360490.7 | c.160+3_160+6del | SDV |  | 1/152052 | 0.99 |
| 3-155084328-G-T |  | ENST00000360490.7 | c.160+1G>T | SDV |  | 9/1461810 | 1 |
| 3-155084329-T-A | rs1446583831 | ENST00000360490.7 | c.160+2T>A | SDV |  | 7/1613958 | 1 |
| 3-155085057-A-G |  | ENST00000360490.7 | c.161-2A>G | SAV |  | 1/1433184 | 0.99 |
| 3-155085094-GGTA-G | rs763866586 | ENST00000360490.7 | c.196+1_196+3del | SDV |  | 1/1419544 | 1 |
| 3-155105123-A-G | rs1717582655 | ENST00000360490.7 | c.197-9871A>G | IV |  | 1/152134 | 0.81 |
| 3-155115156-G-A | rs1718495651 | ENST00000360490.7 | c.358+1G>A | SDV |  | 1/152154 | 1 |
| 3-155116473-TTTCAGATGTCC-T |  | ENST00000360490.7 | c.359-1_368del | SAV |  | 1/814454 | 1 |
| 3-155116478-G-T |  | ENST00000360490.7 | c.359-1G>T | SAV |  | 1/819878 | 1 |
| 3-155116560-G-C | rs1281943901 | ENST00000360490.7 | c.439+1G>C | SDV |  | 3/1586350 | 1 |
| 3-155116560-G-A | rs1281943901 | ENST00000360490.7 | c.439+1G>A | SDV |  | 1/150608 | 1 |
| 3-155116561-T-A | rs1057519024 | ENST00000360490.7 | c.439+2T>A | SDV | P | 3/1436714 | 1 |
| 3-155116564-G-C | rs1053060665 | ENST00000360490.7 | c.439+5G>C | IV |  | 1/1426974 | 0.86 |
| 3-155116662-A-C | rs200435950 | ENST00000360490.7 | c.440-2A>C | SAV | P/LP | 23/1609404 | 1 |
| 3-155116663-G-C | rs781368086 | ENST00000360490.7 | c.440-1G>C | SAV |  | 3/778078 | 1 |
| 3-155116753-AAATATGG-A |  | ENST00000360490.7 | c.530_535+1del | SDV |  | 4/832764 | 1 |
| 3-155116866-A-T |  | ENST00000360490.7 | c.536-2A>T | SAV |  | 1/807770 | 0.99 |
| 3-155116866-A-G |  | ENST00000360490.7 | c.536-2A>G | SAV |  | 1/807772 | 0.98 |
| 3-155116867-G-T | rs759072209 | ENST00000360490.7 | c.536-1G>T | SAV |  | 1/1439738 | 0.99 |
| 3-155116867-G-A | rs759072209 | ENST00000360490.7 | c.536-1G>A | SAV | P/LP | 2/1591738 | 1 |
| 3-155116958-A-G |  | ENST00000360490.7 | p.Asp209Gly | MV |  | 3/1443478 | 0.99 |
| 3-155116982-T-G | rs1718677283 | ENST00000360490.7 | p.Ile217Ser | MV |  | 2/1367902 | 0.93 |
| 3-155116987-G-A | rs1057519023 | ENST00000360490.7 | c.654+1G>A | SDV | P | 2/1316506 | 1 |
| 3-155116988-T-A |  | ENST00000360490.7 | c.654+2T>A | SDV |  | 1/689260 | 1 |
| 3-155116991-G-A |  | ENST00000360490.7 | c.654+5G>A | IV |  | 1/626820 | 0.82 |
| 3-155118744-A-G | rs765591205 | ENST00000360490.7 | c.655-2A>G | SAV | P | 13/1576250 | 0.98 |
| 3-155118745-G-C | rs1718836921 | ENST00000360490.7 | c.655-1G>C | SAV |  | 3/1581874 | 0.98 |
| 3-155118812-G-A | rs199987860 | ENST00000360490.7 | c.720+1G>A | SDV | LP | 14/1577218 | 0.99 |
| 3-155138226-A-T |  | ENST00000360490.7 | p.Glu282Val | MV |  | 1/628246 | 0.9 |
| 3-155140183-A-G |  | ENST00000360490.7 | c.856-8A>G | SRV |  | 1/1440252 | 0.94 |
| 3-155140184-T-G |  | ENST00000360490.7 | c.856-7T>G | SRV |  | 1/1443050 | 0.9 |
| 3-155140187-ATAGGC-A |  | ENST00000360490.7 | c.856-1_859del | SAV |  | 1/1448138 | 0.99 |
| 3-155140188-T-G |  | ENST00000360490.7 | c.856-3T>G | SRV |  | 1/1448374 | 0.94 |
| 3-155140189-A-G | rs770600970 | ENST00000360490.7 | c.856-2A>G | SAV |  | 1/623768 | 0.98 |
| 3-155140291-AG-A | rs776678738 | ENST00000360490.7 | c.957+1del | FS |  | 3/1444918 | 1 |
| 3-155140293-G-C |  | ENST00000360490.7 | c.957+1G>C | SDV |  | 1/1435824 | 1 |
| 3-155141980-CTTTTTTCCAGCCATTCAGCTGGTTGAA-C | rs1452775986 | ENST00000360490.7 | c.958-7_977del | SAV |  | 1/152122 | 0.96 |
| 3-155141988-C-G | rs1468691508 | ENST00000360490.7 | c.958-3C>G | SRV | VUS | 1/833102 | 0.88 |
| 3-155142127-G-T | rs1468741223 | ENST00000360490.7 | p.Arg365Ile | MV |  | 1/152116 | 0.9 |
| 3-155142132-G-T | rs1048105813 | ENST00000360490.7 | c.1094+5G>T | IV | VUS | 7/1461506 | 0.88 |
| 3-155142234-CAG-C | rs760853459 | ENST00000360490.7 | c.1096_1097del | SAV |  | 2/628292 | 0.88 |
| 3-155142330-G-A | rs1721174222 | ENST00000360490.7 | p.Lys396Lys | SRV | VUS | 4/1459430 | 0.94 |
| 3-155142331-G-A |  | ENST00000360490.7 | c.1188+1G>A | SDV | LP | 6/1459320 | 0.95 |
| 3-155142758-A-G | rs61758195 | ENST00000360490.7 | c.1188+428A>G | IV |  | 10/152120 | 0.97 |
| 3-155143438-C-A |  | ENST00000360490.7 | c.1189-5C>A | SRV |  | 3/1459004 | 0.85 |
| 3-155143441-A-G | rs1328070238 | ENST00000360490.7 | c.1189-2A>G | SAV |  | 15/1459150 | 0.99 |
| 3-155143505-G-T |  | ENST00000360490.7 | p.Gly417Gly | SynV |  | 1/1460628 | 0.83 |
| 3-155143560-A-G |  | ENST00000360490.7 | p.Ser436Gly | MV |  | 4/833110 | 0.86 |
| 3-155143568-TGTG-T |  | ENST00000360490.7 | c.1317_1317+2del | InFD |  | 1/1459496 | 0.99 |
| 3-155143571-GGTAAT-G | rs1721287804 | ENST00000360490.7 | c.1317+3_1317+7del | SDV |  | 12/1611258 | 0.97 |
| 3-155143572-G-A | rs776999850 | ENST00000360490.7 | c.1317+1G>A | SDV |  | 3/626148 | 0.97 |
| 3-155143573-T-G | rs752646458 | ENST00000360490.7 | c.1317+2T>G | SDV |  | 17/1459200 | 0.99 |
| 3-155144358-G-C |  | ENST00000360490.7 | c.1318-1G>C | SAV | LP | 1/1452170 | 0.99 |
| 3-155147225-G-C | rs1395068713 | ENST00000360490.7 | c.1497+1G>C | SDV | LP | 8/1574252 | 1 |
| 3-155147225-G-A |  | ENST00000360490.7 | c.1497+1G>A | SDV |  | 3/1422164 | 1 |
| 3-155147226-TA-T |  | ENST00000360490.7 | c.1497+4del | IV |  | 1/776648 | 0.95 |
| 3-155148544-A-G |  | ENST00000360490.7 | c.1498-6A>G | SRV |  | 1/1418544 | 1 |
| 3-155148547-C-G | rs1417837337 | ENST00000360490.7 | c.1498-3C>G | SRV | VUS | 1/152038 | 1 |
| 3-155148548-A-G | rs1721692909 | ENST00000360490.7 | c.1498-2A>G | SAV |  | 1/152204 | 1 |
| 3-155148549-G-A |  | ENST00000360490.7 | c.1498-1G>A | SAV |  | 1/1432532 | 0.99 |
| 3-155148549-G-T | rs1467339413 | ENST00000360490.7 | c.1498-1G>T | SAV |  | 1/1432532 | 0.99 |
| 3-155148650-A-G | rs771842754 | ENST00000360490.7 | p.Asp533Gly | MV | VUS | 7/1451868 | 1 |
| 3-155148654-G-A |  | ENST00000360490.7 | c.1601+1G>A | SDV |  | 1/1448640 | 1 |
| 3-155148654-G-C |  | ENST00000360490.7 | c.1601+1G>C | SDV |  | 1/1448640 | 1 |
| 3-155148655-T-G | rs1403042622 | ENST00000360490.7 | c.1601+2T>G | SDV |  | 2/1599674 | 0.99 |
| 3-155148655-T-C | rs1403042622 | ENST00000360490.7 | c.1601+2T>C | SDV |  | 4/1447714 | 0.92 |
| 3-155148658-G-A | rs201612792 | ENST00000360490.7 | c.1601+5G>A | IV | VUS | 6/1594054 | 0.87 |
| 3-155160445-A-G | rs1179679279 | ENST00000360490.7 | p.Ile553Val | MV |  | 2/1437624 | 1 |
| 3-155160448-G-T |  | ENST00000360490.7 | p.Val554Phe | MV |  | 1/1432506 | 0.86 |
| 3-155166899-TA-T |  | ENST00000360490.7 | c.1661-2del | SAV |  | 1/1461344 | 1 |
| 3-155166900-A-G | rs1723135630 | ENST00000360490.7 | c.1661-2A>G | SAV |  | 2/628260 | 1 |
| 3-155167022-G-A |  | ENST00000360490.7 | c.1780+1G>A | SDV |  | 1/1461226 | 0.98 |
| 3-155167024-A-G | rs1299522334 | ENST00000360490.7 | c.1780+3A>G | IV | VUS | 3/780452 | 0.87 |
| 3-155168490-A-G | rs765231758 | ENST00000360490.7 | c.1781-2A>G | SAV | P/LP | 21/1610110 | 0.89 |
| 3-155168798-G-A |  | ENST00000360490.7 | c.1980+1G>A | SDV |  | 1/827848 | 0.99 |
| 3-155172105-T-G |  | ENST00000360490.7 | c.1981-12T>G | IV |  | 2/1331240 | 0.99 |
| 3-155172110-T-A |  | ENST00000360490.7 | c.1981-7T>A | SRV |  | 1/1372884 | 0.99 |
| 3-155172115-A-G | rs746419069 | ENST00000360490.7 | c.1981-2A>G | SAV |  | 1/1399262 | 0.95 |
| 3-155172115-A-T | rs746419069 | ENST00000360490.7 | c.1981-2A>T | SAV |  | 1/1399260 | 0.95 |
| 3-155172211-A-G | rs758858771 | ENST00000360490.7 | p.Gln692Arg | MV | VUS | 10/1439934 | 0.82 |
| 3-155172213-G-A | rs1170321638 | ENST00000360490.7 | c.2076+1G>A | SDV |  | 1/1435300 | 0.98 |
| 3-155172215-A-G | rs1339057934 | ENST00000360490.7 | c.2076+3A>G | IV | VUS | 4/1430748 | 0.95 |
| 3-155180350-T-A |  | ENST00000360490.7 | c.2154-10T>A | IV |  | 1/830230 | 0.85 |
| 3-155180352-TTTCAAAGGATTA-T | rs1712962174 | ENST00000360490.7 | c.2154-5_2160del | SAV | VUS | 7/628050 | 0.95 |

1.	Høyer H, Hilmarsen HT, Sunder-Plassmann R, et al. A polymorphic AT-repeat causes frequent allele dropout for an *MME* mutational hotspot exon. *J Med Genet*. 2022;59(10):1024-1026. doi:10.1136/jmedgenet-2021-108281

2.	Chen S, Francioli LC, Goodrich JK, et al. A genome-wide mutational constraint map quantified from variation in 76,156 human genomes. *bioRxiv*. Published online 2022:2022.03.20.485034-2022.03.20.485034. doi:10.1101/2022.03.20.485034
